# Impact of pharmacist board certification on health outcomes of critically ill patients: An analysis of the Optimizing Pharmacist-Team Integration for ICU patient Management (OPTIM) study

**DOI:** 10.64898/2026.05.26.26353672

**Authors:** Susan E. Smith, Kelli Henry, Mojdeh S. Heavner, Chelsea Keedy, Huong Duong, Zhetao Chen, XianYan Chen, OPTIM Investigator Team, Andrea Sikora

## Abstract

**BACKGROUND:** Critical care pharmacists (CCPs) reduce adverse drug events (ADEs) and mortality in the intensive care unit (ICU). Board certification is the established professional standard for CCPs but its impact on ICU patient outcomes, including its relationship between CCP characteristics and workload, remain unclear. The purpose of this study was to evaluate the association between pharmacist board certification, CCP workload characteristics, and patient outcomes.

**METHODS:** This was a pre-planned analysis of the multicenter, observational Optimizing Pharmacist-Team Integration for ICU Patient Management (OPTIM) study, including adult ICU patients cared for by CCPs. Patients cared for exclusively by board-certified pharmacists on every ICU day were categorized as the BCP group; those with at least one day of care from a non-board-certified pharmacist comprised the non-BCP group. The primary outcome was hospital mortality; secondary outcomes included the hazard of discharge alive (HDA) from the ICU and hospital. Multivariable logistic regression was used to evaluate the association between BCP and mortality; Fine-Gray competing risk models were used to assess the relationship between BCP and ICU and hospital HDA.

**RESULTS:** A total of 201 pharmacists (184 BCPs; 17 non-BCPs) from 63 institutions caring for 20,537 ICU patients were included. Care provided exclusively by a BCP (vs. ≥ 1 day by a non-BCP) was associated with lower mortality (OR 0.80, 95% CI 0.69–0.92, p=0.002) and both a higher ICU HDA (HR 1.08, 95% CI 1.03–1.13, p<0.001) and hospital HDA (HR 1.19, 95% CI 1.13–1.26, p<0.001).

**CONCLUSION:** Daily ICU care delivered by pharmacists with board certification was independently associated with reduced mortality and improved hazard of discharge alive from the ICU. Board-certified pharmacists may enhance the quality and/or efficiency of critical care pharmacy services. These findings support the role of board certification as a modifiable factor to improve patient outcomes and optimize workload in the ICU.

**Importance and Relevance:** Critical care pharmacists (CCPs), through their vital role in the intensive care unit multi-professional care team, reduce adverse drug events and mortality. However, CCPs remain underutilized therefore limiting care quality. This study, the first to evaluate the impact of pharmacist board certification on patient outcomes, demonstrates that CCP board certification is a modifiable factor associated with reduced mortality and a greater likelihood of successful ICU hospitalization discharge. These results highlight the value of pharmacist board certification to advance critical care pharmacy services, guide workforce development and efficiency of resource utilization, and inform institutional policy for better patient-centered care and outcome-driven performance.

## BACKGROUND

The intensive care unit (ICU) is a high-acuity, high-stakes healthcare environment that relies on an interdisciplinary, team-based approach to provide optimal patient care. Critical care pharmacists (CCPs) are crucial team members in reducing adverse drug events (ADEs) and patient mortality.^1,2^ Studies have shown that over 1.5 million American adults admitted to the ICU each year experience a life-threatening ADE, highlighting the importance of having a medication expert on the care team.^3,4^ Despite their positive impact on patient outcomes, CCPs remain underutilized. Clinical pharmacy services are often limited during evenings and weekends, and CCPs are not available in one-third of U.S. ICUs.^5-7^

The complex and fast-moving ICU environment requiring time-sensitive decision-making, sometimes in the setting of incomplete information, makes ICU healthcare members, including CCPs prone to burnout and other workload-related challenges.^8,9^ Excessive CCP workload may reduce quality of care and worsen patient outcomes. The factors that impact workload and subsequent healthcare worker well-being are multifaceted and constantly evolving.^10^ For CCPs, these can include an excessive patient load and high illness severity that may lead to an increase in workload and hours worked and less than optimal engagement with the rest of the interprofessional ICU team. Beyond these job-related elements, CCP qualifications and training may also affect their workload and outcomes of their patients.

Pharmacists can seek board certification to advance their skills and knowledge in various specialty areas, including critical care, through the Board of Pharmacy Specialties (BPS).^11^ Pharmacist board certification is available to those who have graduated from a recognized school or college of pharmacy (usually with a Doctor of Pharmacy (PharmD) or Bachelor of Science in Pharmacy (BSPharm) degree) who have demonstrated extensive practice experience in their field, passed the standardized board exam, and maintain professional development annually. Board certification provides a structured framework for evidence of competence, quality assurance, professional development, and clinical expertise to stakeholders. Evidence suggests that board certification may lead to improved practice behaviors and patient outcomes. For example, oncology pharmacy board certification has been associated with an increased rate of appropriate chemotherapy regimens in Japan.^12^ Similarly, the Added Qualification in Cardiology (AQCV) has been associated with higher rates of adherence to quality measures in patients with heart failure and myocardial infarction, indicating implications for both patient and financial outcomes.^13^ However, there is limited evidence evaluating the impact of board certification in the ICU setting.

This study aimed to explore the relationship between board certification, CCP workload characteristics, and patient outcomes in the ICU. The primary hypothesis was that board certification would enhance pharmacy services and be associated with reduced mortality and ICU throughput. Ultimately, this study sought to identify potential modifying factors that support CCP practice optimization and advance quality of care.

## METHODS

### Study Design

This multicenter observational study was a pre-planned analysis of the Optimizing Pharmacist-Team Integration for ICU patient Management (OPTIM) database. Full study details have been published previously.^14,15^ In brief, OPTIM was a 64-center study that included 213 CCPs and 33,464 ICU patients. CCPs were recruited from national pharmacy organization listservs and completed study onboarding prior to beginning data collection. Participating CCPs submitted data on their personal demographics, institutional demographics, and characteristics of the ICU team with which they work. Each CCP then prospectively collected daily workload data and enrolled all ICU patients they were assigned to care for for 100 consecutive days. The 100-day periods were staggered across study sites, with data entry occurring between August 2023 and August 2024. Patient characteristics and outcomes data were collected retrospectively from the electronic health record (EHR). All data was input into a secure Research Electronic Data Capture (REDCap) form with patient identifiers removed.^16^ Data points included institutional characteristics, CCP characteristics, ICU practice model characteristics, daily workload and census metrics, and patient characteristics and outcomes. As described in Table 1, novel workload-related variables were calculated including a patient-specific CCP-to-ICU patient ratio and a patient-specific nurse-to-patient ratio. The study was approved by the University of [XXX] Institutional Review Board (PROJECT00011467, April 11, 2023) with a waiver of informed consent for ICU patient enrollment and a partial waiver of consent for pharmacist enrollment. All institutions completed a data use agreement and IRB per institutional requirements. Study procedures adhered to the ethical principles outlined in the Helsinki Declaration of 1975.^14,17^

**Table 1:** Definitions of Patient-Specific Workload Variables Calculated from OPTIM Data.

| Calculated Variable | Description |
| --- | --- |
| Patient-specific CCP-to-ICU patient ratio | If a patient had a 3-day ICU length of stay and the CCP caring for them covered 12, 8, and 14 ICU patients on those three days, then the patient-specific CCP-to-patient ratio would be: $(12+8+14)/3 = 11.3$ (ratio: 1:11). Note that a different CCP may have cared for the patient on different days. |
| Patient-specific nurse-to-patient ratio | If a patient had a 5-day ICU length of stay and nurse caring for them was responsible for 2, 2, 1, 1, and 2 patients on those five days, then the patient-specific nurse-to-patient ratio would be: $(2+2+1+1+2)/5 = 1.6$ (ratio 1: 1.6). |

CCP: Critical care pharmacist; ICU: Intensive care unit

### Study Population

Pharmacists were eligible to participate if they were not enrolled in a training program and an ICU was the primary setting where they cared for patients during weekday day shifts. Multiple pharmacists from the same institution were eligible to participate. Pharmacists were classified as board-certified pharmacist (BCP) or non-board-certified pharmacists (non-BCP) based on the presence of at least one active certification from the Board of Pharmacy Specialties.

A total of 33,464 patients admitted to the ICU and managed by participating CCPs were initially enrolled in the OPTIM REDCap database. For the present analysis, one site was excluded based on stipulations from the data use agreement in the original OPTIM study. Additional patients were excluded according to the following criteria: (1) ICU length of stay (LOS) less than 24 hours, (2) age less than 18 years, (3) lack of continuous pharmacist coverage throughout the ICU stay and (4) missing mortality data.

### Study Outcomes

The primary outcome was hospital mortality, defined as death during the acute hospitalization. Secondary outcomes included the hazard of discharge alive (HDA) from the ICU and the hospital. To examine the impact of board certification, patients were divided into two groups based on whether they received comprehensive medication management (CMM) by a board-certified pharmacist during every day of their ICU stay (BCP group) or whether they received CMM from a pharmacist without board certification on at least one of their ICU days (non-BCP group).

### Study Variables

Collected variables included individual CCP characteristics (e.g., degree(s) earned, active board certification(s), years since terminal training), institutional demographics of each participating site (e.g., geographic region, institution type, pharmacy department staffing model), and characteristics of the interprofessional ICU team (e.g., which professions participate in daily rounds). Daily CCP workload variables included relevant staffing and workload indicators such as the number of interprofessional rounding teams assigned to the pharmacist, number of ICU and non-ICU patients assigned, and number of learners assigned to the pharmacist. Daily patient variables collected for each day of their ICU admission included the pharmacist-to-patient ratio and nurse-to-patient ratio for that specific patient, whether the patient received CMM, and the composition of the medical team caring for the patient (i.e., attending physician, resident(s), fellow(s), advanced practice providers). CMM delivered on interprofessional rounds was defined as pharmacist participation in multidisciplinary rounds and provision of CMM, including comprehensive medication review, development and implementation of individualized medication therapy recommendations, and, where authorized, initiation, modification, or prescribing of medications under collaborative agreements or protocols.^18^ Additional patient variables collected retrospectively included baseline demographics (e.g., age, sex, race/ethnicity), severity of illness scores (i.e., sequential organ failure assessment [SOFA] and medication regimen complexity in the ICU [MRC-ICU] during the first 24 hours of ICU admission), number of days admitted to the ICU and the hospital, and mortality.

### Statistical Methodology

Descriptive statistics were used to report characteristics of institutions, CCPs, workload, patients, and patient outcomes. Multivariable logistic regression and Fine-Gray competing risk models were applied to examine factors associated with patient mortality and HDA, respectively. A predefined set of variables were selected for inclusion in the regression analyses. The primary exposure variable was BCP, defined as whether the patient was cared for exclusively by BCPs during their ICU stay. The secondary exposure variable was the average total number of ICU patients cared for (CCP-to-ICU patient ratio). Additional covariates included age, sex, SOFA score during the first 24 hours of ICU stay, MRC-ICU score during the first 24 hours of ICU stay,^19^ ICU admission day of the week, ICU type, hospital type, nurse-to-patient ratio, coverage from a PGY2 or fellowship trained pharmacist every day of ICU admission, and CMM delivered on interprofessional rounds. Multicollinearity was assessed among related predictors using variance inflation factors (VIFs) and pairwise correlation matrices. A threshold of VIF>5 or r≥0.7 was considered indicative of potential multicollinearity. A complete-case approach was applied, restricting the regression models to patients with non-missing data for the outcomes and all selected covariates. The same exposure variables and covariates were included in Fine-Gray competing risks models to evaluate ICU and hospital HDA, with mortality as a competing event. All analyses were performed with R, version 4.3.0, with a two-side p-value less than 0.05 considered statistically significant.

## RESULTS

### Characteristics of institutions, pharmacists, workload, and patients

A total of 201 pharmacists from 63 institutions and 20,537 patients were included in this study (Figure 1).

**Figure 1:**
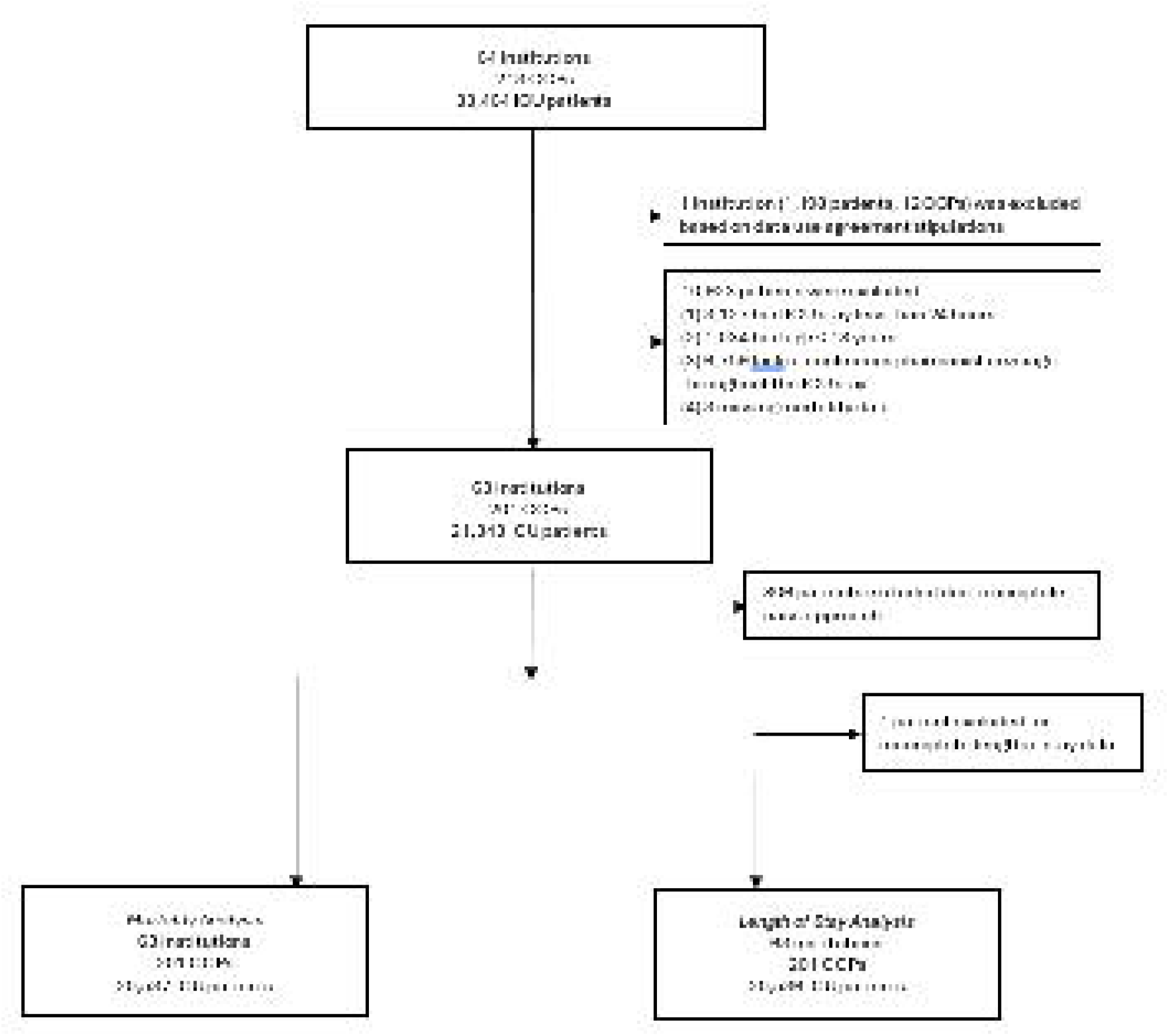
Patient and Pharmacist Inclusion Flow Diagram.

#### Institutional characteristics

Details on institutional characteristics are summarized in Table 2. The 63 institutions represented all geographic regions of the U.S. The majority were academic medical centers (60.3%), followed by community teaching hospitals (25.4%). Nearly half of participating institutions were large hospitals with more than 600 hospital beds and more than 100 ICU beds. Half of institutions were designated level I adult trauma centers and 62% were comprehensive stroke centers. There were a variety of ICU types represented in the study, with medical ICU being the most common, followed by mixed medical/surgical ICU and surgical/trauma ICU.

**Table 2:** Institutional and Pharmacist Characteristics.

| Institutional Characteristics (N=63) |  | Pharmacist Characteristics (N=201) |  |  |  |
| --- | --- | --- | --- | --- | --- |
| Variable | Number (%) | Variable | Number (%) | BCP (n=184) | Non-BCP (n=17) |
| **Institution type** |  | **Degree(s) Earned** |  |  |  |
| Academic Medical Center | 38 (60.3) | Doctor of Pharmacy | 199 (99.0) | 182 (97.8) | 17 (100) |
| Community Teaching Hospital | 16 (25.4) | Bachelor of Science in Pharmacy | 22 (10.9) | 21 (11.4) | 1 (5.9) |
| Community Non-Teaching Hospital | 8 (12.7) | Master of Science | 15 (7.5) | 13 (7.1) | 2 (11.8) |
| Government/VA/Military | 1 (1.6) | Doctor of Philosophy | 1 (0.5) | 1 (0.5) | 0 (0) |
| **Region** |  | **Years since graduation from training** |  |  |  |
| Northeast | 10 (15.9) | Years in practice, mean (SD) | 8.1 (6.8) | 8.3 (6.7) | 6.4 (8.3) |
| Southeast | 20 (31.8) | **Primary ICU Type** |  |  |  |
| Midwest | 16 (25.4) | Medical | 57 (28.4) | 52 (28.2) | 5 (29.4) |
| Northwest | 4 (6.3) | Surgical/Trauma | 23 (11.4) | 20 (10.9) | 3 (17.6) |
| Southwest | 11 (17.5) | Surgical | 11 (5.5) | 11 (6.0) | 0 (0) |
| International | 2 (3.2) | Cardiothoracic | 16 (8.0) | 13 (7.1) | 3 (17.6) |
| **Highest Level of Stroke Designation** |  | Cardiac | 11 (5.5) | 11 (6.0) | 0 (0) |
| Comprehensive Stroke Center | 39 (61.9) | Neurosurgery/Neurology | 14 (7.0) | 13 (7.1) | 1 (5.9) |
| Thrombectomy-Capable Stroke Center | 4 (6.4) | Pediatric | 4 (2.0) | 4 (2.2) | 0 (0) |
| Primary Stroke Center | 16 (25.4) | Mixed Medical-Surgical | 33 (16.4) | 30 (16.3) | 3 (17.6) |
| Acute Stroke-Ready Hospital | 1 (1.6) | Burn | 3 (1.5) | 3 (1.6) | 0 (0) |
| No Stroke Certifications | 3 (4.8) | No most common ICU | 14 (7.0) | 13 (7.1) | 1 (5.9) |
| **Number of Hospital Beds** |  | Other | 15 (7.5) | 14 (7.6) | 1 (5.9) |
| <200 | 2 (3.2) | **Highest Level of Postgraduate Training** |  |  |  |
| 200-400 | 17 (27.0) | Post-graduate Year 1 (PGY1) | 42 (20.9) | 38 (20.7) | 6 (35.3) |
| 401-600 | 14 (22.2) | Post-graduate Year 2 (PGY2) | 136 (67.7) | 128 (69.6) | 8 (47.1) |
| >600 | 30 (47.6) | Fellowship | 3 (1.5) | 3 (1.6) | 0 (0) |
| **Number of ICU Beds** |  | No postgraduate training | 17 (8.5) | 14 (7.6) | 3 (17.6) |
| <26 | 6 (9.5) | **Active Board Certification(s)** |  |  |  |
| 26-50 | 13 (20.6) | BCPS | 74 (36.8) | 74 (40.2) | 0 (0) |
| 51-75 | 9 (14.3) | BCCCP | 150 (74.6) | 150 (81.5) | 0 (0) |
| 76-100 | 6 (9.5) | BCCP | 7 (3.5) | 7 (3.8) | 0 (0) |
| >100 | 29 (46.1) | BCPPS | 3 (1.5) | 3 (1.6) | 0 (0) |
| **Adult Trauma Center Type** |  | BCEMP | 1 (0.5) | 1 (0.5) | 0 (0) |
| Level I | 32 (50.8) | Other board certification | 4 (2.0) | 4 (2.2) | 0 (0) |
| Level II | 14 (22.2) | No active board certification | 17 (8.5) | 0 (0) | 17 (100) |
| Level III | 6 (9.5) |  |  |  |  |
| Level IV | 1 (1.6) |  |  |  |  |
| N/A | 10 (15.9) |  |  |  |  |

VA: Veterans Affairs; ICU: Intensive care unit; BCPS: Board Certified Pharmacotherapy Specialist; BCCCP: Board Certified Critical Care Pharmacist; BCCP: Board Certified Cardiology Pharmacist; BCPPS: Pediatric Pharmacy Specialty Certification; BCEMP: Board Certified Emergency Medicine Pharmacist; N/A: Not applicable; SD: standard deviation
Non-BCP and BCP represent the total number of pharmacists without or with board certification, respectively.

#### Pharmacist characteristics

Details on CCP characteristics are summarized in Table 2. Of the respondents, 99.0% held a Doctor of Pharmacy (PharmD) degree and 67.7% reported post-graduate year 2 (PGY-2) training as the highest level of postgraduate education. Most respondents (91.5%) held at least one board certification, with board-certified critical care pharmacist (BCCCP) being the most common credential (74.6%) and board-certified pharmacotherapy specialist (BCPS) as the next most frequent (36.8%). Respondents had a mean of 8.1 (standard deviation [SD] 6.8) years of experience since completing terminal training. Patients in the BCP group were cared for by CCPs with more years of experience than those in the non-BCP group (mean 8.3 [SD 6.7] vs 6.4 [SD 8.3] years) (Table 3).

**Table 3:**
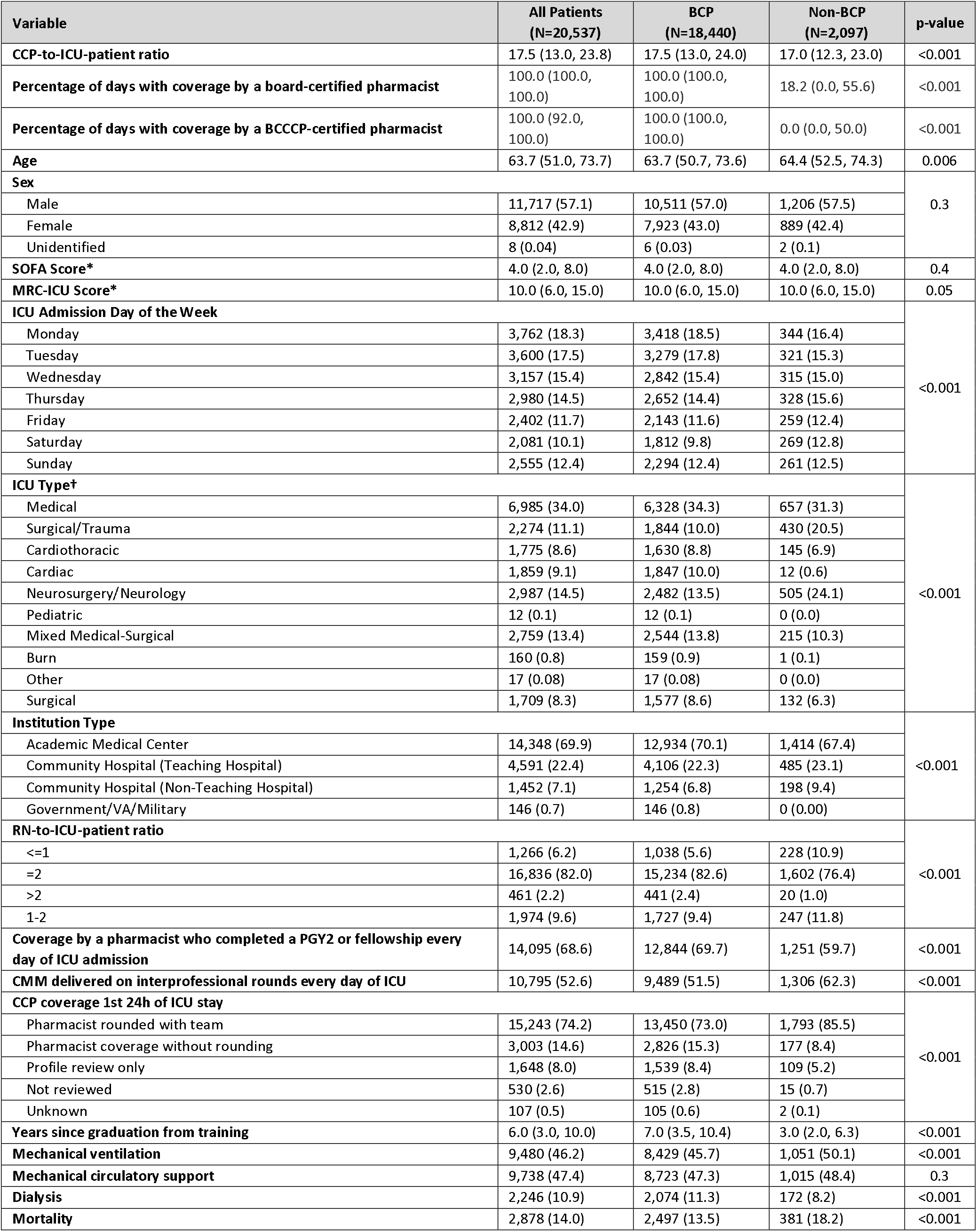
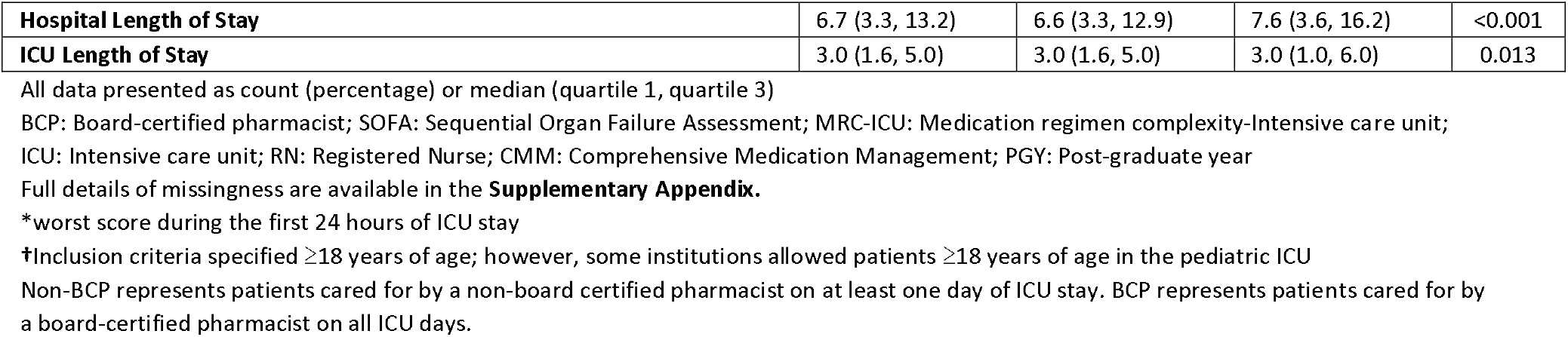
Patient Characteristics (N=20,537)

#### Patient characteristics

Details on patient characteristics are summarized in Table 3. The median age of patients included in the analysis was 63.7 years (IQR 51.0-73.7) and 57.1% were male. Baseline illness severity was moderate, with a median SOFA score of 4 (IQR 2-8) and MRC-ICU score of 10 (IQR 6-15). During hospitalization, 46.2% of patients required mechanical ventilation and 47.4% required mechanical circulatory support. Renal replacement therapy was initiated in 10.9% of patients. There were no significant differences in severity of illness or the need for mechanical circulatory support between patients in the BCP and non-BCP groups. However, patients in the BCP group required mechanical ventilation less frequently (45.7% vs 50.1%, p<0.001) and dialysis more frequently (11.3% vs 8.2%, p<0.001) when compared with patients in the non-BCP group.

#### Workload

Workload metrics, calculated at the patient level and compared between BCP and non-BCP groups, are summarized in Table 3. The median patient-specific CCP-to-ICU patient ratio was 17.5 (IQR 13.0-23.8). During the first 24 hours of ICU admission, 74.2% of patients were rounded on by a CCP as part of a multidisciplinary team. Fewer patients in the BCP group were rounded on during the first 24 hours of ICU admission than in the non-BCP group (73.0% vs 85.5%, p<0.001). Similarly, less CMM was performed during interprofessional rounds in the BCP group (51.5% vs 62.3%, p<0.001).

### Patient outcomes

The overall mortality rate was 14.0%. The median ICU LOS was 3 days (IQR 1.6-5.0) and the median hospital LOS was 6.7 days (IQR 3.3-13.2). Mortality was lower among patients managed by BCPs every day of their ICU stay (13.5% vs 18.2%, p<0.001) (Table 3). In multivariable logistic regression analysis, patients cared for by BCPs every day had 20% lower odds of death compared with those with at least one day of non-BCP care (OR 0.80, 95% CI: 0.69–0.92, p=0.002) (Table 4).

**Table 4.**
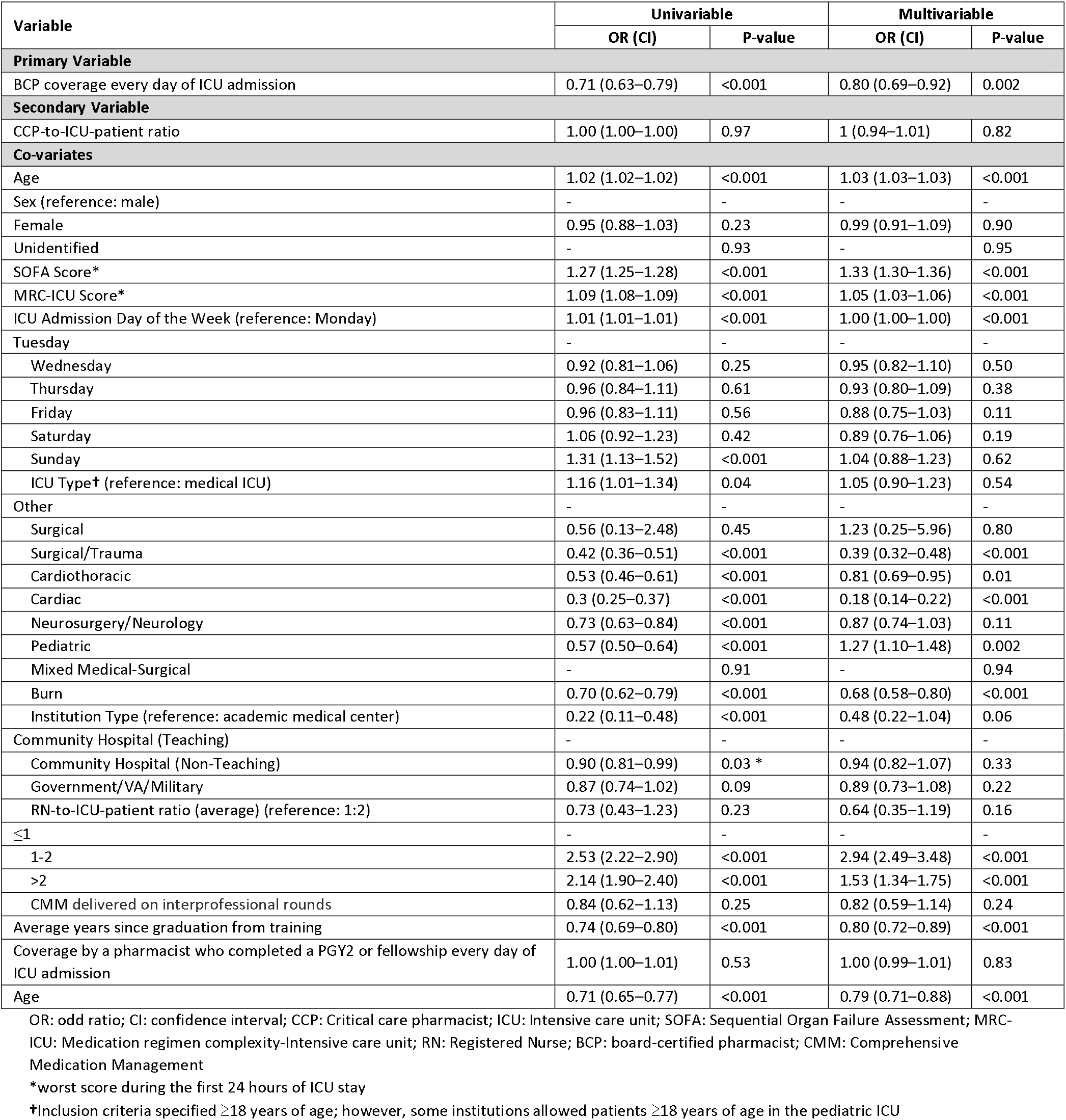
Univariable and Multivariable Logistic Regression Analyses for Mortality Based on Complete Cases Data (N = 20,537)

| Variable | Univariable |  | Multivariable |  |
| --- | --- | --- | --- | --- |
|  | OR (CI) | P-value | OR (CI) | P-value |
| **Primary Variable** |  |  |  |  |
| BCP coverage every day of ICU admission | 0.71 (0.63–0.79) | <0.001 | 0.80 (0.69–0.92) | 0.002 |
| **Secondary Variable** |  |  |  |  |
| CCP-to-ICU-patient ratio | 1.00 (1.00–1.00) | 0.97 | 1 (0.94–1.01) | 0.82 |
| **Co-variates** |  |  |  |  |
| Age | 1.02 (1.02–1.02) | <0.001 | 1.03 (1.03–1.03) | <0.001 |
| Sex (reference: male) | - | - | - | - |
| Female | 0.95 (0.88–1.03) | 0.23 | 0.99 (0.91–1.09) | 0.90 |
| Unidentified | - | 0.93 | - | 0.95 |
| SOFA Score* | 1.27 (1.25–1.28) | <0.001 | 1.33 (1.30–1.36) | <0.001 |
| MRC-ICU Score* | 1.09 (1.08–1.09) | <0.001 | 1.05 (1.03–1.06) | <0.001 |
| ICU Admission Day of the Week (reference: Monday) | 1.01 (1.01–1.01) | <0.001 | 1.00 (1.00–1.00) | <0.001 |
| Tuesday | - | - | - | - |
| Wednesday | 0.92 (0.81–1.06) | 0.25 | 0.95 (0.82–1.10) | 0.50 |
| Thursday | 0.96 (0.84–1.11) | 0.61 | 0.93 (0.80–1.09) | 0.38 |
| Friday | 0.96 (0.83–1.11) | 0.56 | 0.88 (0.75–1.03) | 0.11 |
| Saturday | 1.06 (0.92–1.23) | 0.42 | 0.89 (0.76–1.06) | 0.19 |
| Sunday | 1.31 (1.13–1.52) | <0.001 | 1.04 (0.88–1.23) | 0.62 |
| ICU Type† (reference: medical ICU) | 1.16 (1.01–1.34) | 0.04 | 1.05 (0.90–1.23) | 0.54 |
| Other | - | - | - | - |
| Surgical | 0.56 (0.13–2.48) | 0.45 | 1.23 (0.25–5.96) | 0.80 |
| Surgical/Trauma | 0.42 (0.36–0.51) | <0.001 | 0.39 (0.32–0.48) | <0.001 |
| Cardiothoracic | 0.53 (0.46–0.61) | <0.001 | 0.81 (0.69–0.95) | 0.01 |
| Cardiac | 0.3 (0.25–0.37) | <0.001 | 0.18 (0.14–0.22) | <0.001 |
| Neurosurgery/Neurology | 0.73 (0.63–0.84) | <0.001 | 0.87 (0.74–1.03) | 0.11 |
| Pediatric | 0.57 (0.50–0.64) | <0.001 | 1.27 (1.10–1.48) | 0.002 |
| Mixed Medical-Surgical | - | 0.91 | - | 0.94 |
| Burn | 0.70 (0.62–0.79) | <0.001 | 0.68 (0.58–0.80) | <0.001 |
| Institution Type (reference: academic medical center) | 0.22 (0.11–0.48) | <0.001 | 0.48 (0.22–1.04) | 0.06 |
| Community Hospital (Teaching) | - | - | - | - |
| Community Hospital (Non-Teaching) | 0.90 (0.81–0.99) | 0.03 * | 0.94 (0.82–1.07) | 0.33 |
| Government/VA/Military | 0.87 (0.74–1.02) | 0.09 | 0.89 (0.73–1.08) | 0.22 |
| RN-to-ICU-patient ratio (average) (reference: 1:2) | 0.73 (0.43–1.23) | 0.23 | 0.64 (0.35–1.19) | 0.16 |
| ≤1 | - | - | - | - |
| 1-2 | 2.53 (2.22–2.90) | <0.001 | 2.94 (2.49–3.48) | <0.001 |
| >2 | 2.14 (1.90–2.40) | <0.001 | 1.53 (1.34–1.75) | <0.001 |
| CMM delivered on interprofessional rounds | 0.84 (0.62–1.13) | 0.25 | 0.82 (0.59–1.14) | 0.24 |
| Average years since graduation from training | 0.74 (0.69–0.80) | <0.001 | 0.80 (0.72–0.89) | <0.001 |
| Coverage by a pharmacist who completed a PGY2 or fellowship every day of ICU admission | 1.00 (1.00–1.01) | 0.53 | 1.00 (0.99–1.01) | 0.83 |
| Age | 0.71 (0.65–0.77) | <0.001 | 0.79 (0.71–0.88) | <0.001 |

OR: odd ratio; CI: confidence interval; CCP: Critical care pharmacist; ICU: Intensive care unit; SOFA: Sequential Organ Failure Assessment; MRC-ICU: Medication regimen complexity-Intensive care unit; RN: Registered Nurse; BCP: board-certified pharmacist; CMM: Comprehensive Medication Management
\*worst score during the first 24 hours of ICU stay
†Inclusion criteria specified ≥18 years of age; however, some institutions allowed patients ≥18 years of age in the pediatric ICU

Patients managed exclusively by BCPs had shorter hospital LOS compared to those managed at least one day by non-BCPs (median 6.6 [IQR 3.3–12.9] vs 7.56 [IQR 3.6–16.2] days, p<0.001). ICU LOS for patients in the BCP group and those in the non-BCP group were 3.0 days (IQR 1.6–5.0) and 3.0 days (IQR 1.0–6.0), respectively, with p=0.013 (Table 3). In Fine-Gray competing risks models, patients who received care by BCPs every ICU day had higher HDA from the ICU (HR 1.08 [CI 1.03–1.13], p<0.001) and hospital (HR 1.19 [CI 1.13–1.26], p<0.001), accounting for in-ICU and in-hospital mortality as competing events (Tables 5). No evidence of significant multicollinearity was observed among predictors (Supplemental Table 1 and Supplemental Figure 1), with VIFs ranging from 1.03 to 1.57. Pairwise correlation analysis indicated weak correlations between BCP group and years of experiences (r=0.12) and between BCP group and PGY2 or fellowship training (r=0.07).

**Table 5.** Fine-Gray Competing Risks Model for ICU & Hospital Hazard of Discharge Alive with Mortality as a Competing Event (N = 20,536)

| Variable | ICU Hazard of Discharge Alive |  | Hospital Hazard of Discharge Alive |  |
| --- | --- | --- | --- | --- |
|  | Hazard Ratio (CI) | P-value | Hazard Ratio (CI) | P-value |
| **Primary Variable** |  |  |  |  |
| BCP coverage every day of ICU admission | 1.08 (1.03–1.13) | < 0.001 | 1.19 (1.13–1.26) | < 0.001 |
| **Secondary Variable** |  |  |  |  |
| CCP-to-ICU-patient ratio | 1.00 (1.00–1.00) | 0.25 | 1.00 (1.00–1.00) | 0.003 |
| **Co-variables** |  |  |  |  |
| Age | 1.00 (1.00–1.00) | < 0.001 | 0.99 (0.99–0.99) | < 0.001 |
| Sex (reference: male) | - | - | - | - |
| Female | 1.01 (0.97–1.04) | 0.32 | 1.01 (0.98–1.04) | 0.60 |
| Unidentified | 1.62 (1.01–2.61) | 0.05 | 1.39 (0.74–2.63) | 0.31 |
| SOFA Score* | 0.93 (0.93–0.94) | < 0.001 | 0.90 (0.90–0.91) | < 0.001 |
| MRC-ICU Score* | 0.98 (0.98–0.98) | < 0.001 | 0.98 (0.97–0.98) | < 0.001 |
| ICU Admission Day of the Week (reference: Monday) | - | - | - | - |
| Tuesday | 1.02 (0.98–1.07) | 0.35 | 1.04 (0.98–1.09) | 0.20 |
| Wednesday | 1.05 (1.00–1.11) | 0.03 | 1.02 (0.97–1.08) | 0.43 |
| Thursday | 1.05 (1.00–1.11) | 0.05 | 1.03 (0.98–1.09) | 0.25 |
| Friday | 0.89 (0.85–0.94) | < 0.001 | 0.93 (0.87–0.99) | 0.01 |
| Saturday | 0.87 (0.83–0.92) | < 0.001 | 0.93 (0.88–0.99) | 0.03 |
| Sunday | 0.9 (0.93–1.03) | 0.35 | 0.94 (0.89–1.00) | 0.05 |
| ICU Type† (reference: medical ICU) | - | - | - | - |
| Other | 0.90 (0.45–1.77) | 0.75 | 1.13 (0.39–3.25) | 0.83 |
| Surgical | 1.18 (1.12–1.24) | < 0.001 | 1.11 (1.05–1.18) | < 0.001 |
| Surgical/Trauma | 0.97 (0.92–1.02) | 0.21 | 1.01 (0.95–1.06) | 0.85 |
| Cardiothoracic | 1.18 (1.11–1.25) | < 0.001 | 1.45 (1.36–1.55) | < 0.001 |
| Cardiac | 0.99 (0.94–1.05) | 0.76 | 1.15 (1.08–1.24) | < 0.001 |
| Neurosurgery/Neurology | 0.90 (0.86–0.94) | < 0.001 | 0.93 (0.89–0.98) | 0.006 |
| Pediatric | 1.21 (0.69–2.15) | 0.51 | 1.55 (0.82–2.91) | 0.17 |
| Mixed Medical-Surgical | 1.28 (1.22–1.34) | < 0.001 | 1.02 (0.96–1.08) | 0.55 |
| Burn | 0.67 (0.60–0.76) | < 0.001 | 0.94 (0.80–1.09) | 0.40 |
| Institution Type (reference: academic medical center) | - | - | - | - |
| Community Hospital (Teaching) | 1.16 (1.12–1.20) | < 0.001 | 1.01 (0.97–1.06) | 0.62 |
| Community Hospital (Non-Teaching) | 0.80 (0.75–0.85) | < 0.001 | 1.94 (1.77–2.13) | < 0.001 |
| Government/VA/Military | 1.20 (0.97–1.50) | 0.10 | 3.08 (2.24–4.22) | < 0.001 |
| RN-to-ICU-patient ratio (average) (reference: 1:2) | - | - | - | - |
| ≤1 | 0.77 (0.72–0.83) | < 0.001 | 0.69 (0.48–0.74) | < 0.001 |
| 1-2 | 0.72 (0.69–0.75) | < 0.001 | 0.72 (0.68–0.75) | < 0.001 |
| >2 | 1.06 (0.97–1.16) | 0.17 | 0.99 (0.90–1.10) | 0.87 |
| CMM delivered on interprofessional rounds | 1.39 (1.34–1.43) | < 0.001 | 1.35 (1.29–1.40) | < 0.001 |
| Average years since graduation from training | 1.00 (1.00–1.00) | 0.63 | 1.00 (1.00–1.01) | 0.13 |
| Coverage by a pharmacist who completed a PGY2 or fellowship every day of ICU admission | 1.20 (1.15–1.24) | < 0.001 | 1.01 (0.97–1.06) | 0.51 |

CI: Confidence interval; CCP: Critical care pharmacist; ICU: Intensive care unit; SOFA: Sequential Organ Failure Assessment; MRC-ICU: Medication regimen complexity-Intensive care unit; RN: Registered Nurse; BCP: board-certified pharmacist; CMM: Comprehensive Medication Management; PGY: Post-graduate year
\*worst score during the first 24 hours of ICU stay
†Inclusion criteria specified ≥18 years of age; however, some institutions allowed patients ≥18 years of age in the pediatric ICU

## DISCUSSION

In this multicenter observational study, care provided exclusively by BCPs was associated with improved patient outcomes, including lower mortality and higher ICU and hospital HDA. These findings highlight the potential role of pharmacist board certification in addressing an ongoing issue with underutilization of CCPs, a multifaceted concern that may be contributed to by staffing shortages and availability of qualified CCPs. Suboptimal staffing of CCPs in the critical care setting has been associated with excessive workload and negative patient outcomes.^5-9^ Improving pharmacist qualifications through board certification can mitigate these challenges. After adjusting for potential factors affecting patient outcomes, including severity of illness and other patient characteristics, institution types, CMM status, and nurse-to-patient ratio,^20^ patients who received care from BCPs every day of their ICU stay had significantly lower odds of mortality compared with those who received care from a non-BCP on at least one day. Additionally, care provided exclusively by BCPs was associated with improved ICU and hospital HDA when accounting for in-ICU and in-hospital mortality as competing risks, suggesting a positive impact on patient recovery and healthcare utilization.

While patients in the BCP group were cared for by pharmacists with more years of experience than those in the non-BCP group, pairwise correlation analysis indicated only a weak correlation between BCP status and years of experience. Moreover, the number of years of experience was not independently associated with patient outcomes in either the logistic regression or Fine-Gray competing risk models, indicating that board certification may play a distinct independent role beyond experience alone. Additionally, while care by a PGY2 residency and/or fellowship-trained CCP was associated with improved patient outcomes, the relationship between board certification status and patient outcomes was significant after accounting for post-graduate training of CCPs in the multivariable analyses, again suggesting that board certification provides a distinct clinical benefit, though it likely builds synergistically upon the foundation of advanced training. Finally, although BCPs represented a large proportion of the cohort, the substantial number of outcome events in both groups, the adequate statistical power, and the consistency of results across adjusted models mitigate concerns regarding exposure imbalance.

Board certification of clinical pharmacists, endorsed by the American College of Clinical Pharmacy, serves as a validated indicator of specialized pharmacotherapy expertise.^21^ It offers a standardized framework for assessing competence, supporting ongoing professional development, and maintaining clinical proficiency through rigorous evaluation. BCPs acquire specialized pharmacotherapeutic knowledge and demonstrate competency through standardized assessment.^11^ The requirement for ongoing professional development to maintain certification further enhances clinical expertise and ensures BCPs remain up to date with evidence-based practices. These factors may translate into identification of more impactful medication-related problems, contributing to both improved survival and recovery, as reflected by shorter lengths of stay.

This enhanced ability to identify impactful problems may also explain the unexpected differences in workload data. Notably, patients in the BCP group received less frequent pharmacist rounding during the first 24 hours of ICU admission and less CMM during interprofessional rounds compared to those in the non-BCP group, yet they demonstrated better clinical outcomes. While counter-intuitive, this finding may reflect a qualitative difference in the nature of pharmacy services provided rather than a quantitative deficiency in care. BCPs may resolve high-impact medication-related problems more efficiently, reducing the necessity for repeated bedside interventions. This suggests that board certification may enhance the quality and efficiency of clinical pharmacy practice, rather than simply increasing the volume of interventions delivered. In this context, fewer rounding encounters coupled with better clinical outcomes provided by BCPs may reflect more targeted, effective pharmaceutical care rather than reduced engagement. The findings from this study are consistent with prior research suggesting that board certification may enhance the quality of clinical practice. For instance, oncology pharmacy board certification has been linked to greater adherence to guideline-directed chemotherapy.^12^ Likewise, the Added Qualification in Cardiology has been associated with higher adherence to quality measures in patients with heart failure and myocardial infarction.^13^ Collectively, these studies support the value of pharmacy certification in advancing clinical services, bringing benefits to both patient and institutions.

These findings have important implications for workforce development and institutional policy. As healthcare systems increasingly emphasize value-based care and outcome-driven performance, pharmacist board certification may represent a modifiable factor associated with improved patient outcomes. Beyond faster recovery, the association between board certification and higher HDA can lead to reduced hospital-associated complications including nosocomial infections, improved quality of life, and decreased financial burdens for patients.^22^ In addition to patient-centered implications, this association could also reduce staffing burden associated with prolonged ICU stays, improve operational efficiency, and lower both direct and indirect hospitalization costs. Recognition and support of board certification therefore can enhance the quality of critical care pharmacy services, patient-centered outcomes, and resource utilization. While it should be noted that this study included substantial representation from academic medical centers and a high prevalence of board certification, obtaining this credential is a relatively inexpensive endeavor that could likely be supported and prioritized, even in lower resource settings.

This study has several strengths. It is the first multicenter study evaluating the impact of pharmacy board certification on patient outcomes in the critical care setting. The multicenter design and inclusion of a large population of ICU patients enhance the generalizability of the finding across diverse ICU practice environments. Furthermore, the use of objective patient-centered outcomes, such as mortality and ICU and hospital HDA, strengthens the clinical relevance of the results.

Although efforts were made to align methods with a recent ACCP statement on studying the impact of pharmacist board certification,^23^ several limitations should be noted. As an observational study, causal relationships cannot be established, and residual confounding may remain. For example, it is possible that pharmacists have additional certifications (other than board certification) that were not accounted for. Given the voluntary nature of enrollment, selection bias for pharmacists who were willing to commit to a time-intensive data collection process is possible. Additionally, the BCP group had greater years of experience compared to the non-BCP group, which may also be reflective of potential selection bias of those willing to participate in the study. Due to high prevalence of BCPs across participating sites and the observational study design, a direct comparison between patients receiving care exclusively from BCPs and those receiving care exclusively from non-BCPs was not feasible. As a result, outcomes observed in the non-BCP group may still reflect some degree of board-certified pharmacist involvement. Future studies are warranted to further explore the causal relationships between pharmacist board certification, ICU workload, and patient outcomes.

## CONCLUSION

This multicenter observational study is the first to demonstrate the association between pharmacist with board certification and improved patient outcomes in the critical care setting. Care provided by BCPs was associated with lower mortality and higher rate of discharge alive from the ICU and hospital. These findings support the promotion of pharmacist continuous professional development and highlight the value of board certification as a strategy to enhance the quality and impact of critical care pharmacy services, as well as optimize patient outcomes and clinician workload.

## Data Availability

The data that support the findings of this study are available from the corresponding author upon reasonable request.

## Acknowledgements

See the supplementary material for a full list of OPTIM Investigator Team authors and acknowledgements.

## Notes

**Conflicts of interest:** The authors have no conflicts of interest. A list of relevant disclosures is provided in the Supplementary Appendix.

**Funding information:** Funding through the Agency for Healthcare Research and Quality for Drs. Sikora and Smith was provided through R21HS028485 and R01HS029009. Funding through the University of Maryland, Baltimore, Institute for Clinical & Translational Research voucher program was provided to Dr. Heavner. Funding through the ASHP Research and Education Foundation was provided through a research grant to Dr. Heavner. Funding through the Board of Pharmacy Specialties was provided through a research grant to Dr. Smith. Funding through the American College of Clinical Pharmacy Critical Care Practice & Research Network was provided through research grants to Drs. Sikora, Henry, and Murray. Funding from the National Center for Advancing Translational Sciences of the National Institutes of Health under Award Number UL1TR002378 supported the REDCap license. The content is solely the responsibility of the authors and does not necessarily represent the official views of any funders.

**Data sharing:** The data that support the findings of this study are available from the corresponding author upon reasonable request.

### Competing Interest Statement

The authors have declared no competing interest.

### Author Declarations

The study was approved by the University of Georgia Institutional Review Board (PROJECT00011467, April 11, 2023) with a waiver of informed consent for ICU patient enrollment and a partial waiver of consent for pharmacist enrollment. All institutions completed a data use agreement and IRB per institutional requirements.

