## Supplementary Appendix for "Impact of pharmacist board certification on health outcomes of critically ill patients: An analysis of the Optimizing Pharmacist-Team Integration for ICU patient Management (OPTIM) study"

This appendix has been provided by the authors to give readers additional information about the work.

Sarah A.H. Adams, PharmD, Piedmont Athens Regional

Sarah Adie, PharmD, FACC, University of Michigan Health

Maria O. Agunsoye, PharmD, Abbott Northwestern Hospital

May Alanazi, PharmD, Health Holding Company

Abdullah M. Alhammad, PharmD, College of Pharmacy, King Saud University, Riyadh, Saudi Arabia

Enas Alkurdi, PharmD, King Hussein Cancer Center, Jordan

Shannon J. Allcron, PharmD, Owensboro Health Regional Hospital

Christopher A. Anderson, PharmD, Indiana University Health

Whitney S. Anderson, PharmD, Premier Health - Miami Valley Hospital

Eve Anderson, PharmD, Indiana University Academic Health Center

Katherine L. Artman, PharmD, University of Mississippi Medical Center

Wedad Bilal Awad, PharmD, King Hussein Cancer Center

Leslie D. Banuelos, PharmD, University of Maryland Medical Center, Cedars-Sinai Medical Center

Alyson T. Basting, PharmD, Indiana University Health Adult Academic Health Center

Jennifer Bauer, PharmD, OhioHealth Riverside Methodist Hospital

Katherine Beach, PharmD, Atrium Health Wake Forest Baptist

Erin Beauclair, PharmD, Avera McKennan Hospital and University Health Center

Angel Becker, PharmD, BCPS, DPLA, Abbott Northwestern Hospital

Rachel M. Belcher, PharmD, Onvida Health

Amanda Bernarde, PharmD, University of Alabama at Birmingham

Kara L. Birrer, PharmD, FCCP, Orlando Health Orlando Regional Medical Center

Christopher J. Bollinger, PharmD, University of North Carolina

Allison N. Boyd, PharmD, FCCM, Eskenazi Health

Trisha N. Branan, PharmD, FCCM, University of Georgia College of Pharmacy

Jessica M. Brochu, PharmD, Henry Ford Hospital

Michelle Brownstein, PharmD, Cleveland Clinic Weston

Quang V. Bui, PharmD, UCSF Benioff Children's Hospital Oakland

Marisha Burden, MD, MBA, University of Colorado School of Medicine

Simona O. Butler, PharmD, University of Michigan Health

Stacy L. Campbell-Bright, PharmD, UNC

Christina L. Candeloro, PharmD, Roswell Park Comprehensive Cancer Center

Irene Capistrano, PharmD, Orlando Health Bayfront Hospital

Breanna L. Carter, PharmD, Erlanger Health System, University of Tennessee College of Medicine-Department of Surgery

Gianna Lauren Casal, PharmD, Massachusetts General Hospital

Alyssa M. Castillo, PharmD, Texas A&M University Irma Lerma Rangel College of Pharmacy

Aaron M. Chase, PharmD, MUSC

Joshua Chestnutt, PharmD, Piedmont Columbus Regional Midtown

Stephanie R. L. Ciapala, PharmD, Cleveland Clinic

Angela M. Clark, PharmD, Michigan Medicine

Kathryn A. Connor, PharmD, FCCM, St. John Fisher University Wegmans School of Pharmacy

Jessica M. Connor, PharmD, University of Maryland Medical Center

Jenna M. Corvelli, PharmD, University of Rochester Medical Center

Emma Covington, PharmD, University of Virginia

Jenna F. Cox, PharmD, FCCM, Prisma Health Richland Hospital, University of South Carolina College of Pharmacy

Shawnee N. Daniel-McCalla, PharmD, University of Maryland Medical Center

Hannah Davis, PharmD, University of North Carolina Medical Center

Aubrey A. Defayette, PharmD, Roswell Park Comprehensive Cancer Center

John W. Devlin, PharmD, MCCM, Northeastern University School of Pharmacy, Brigham and Women’s Hospital, Division of Pulmonary and Critical Care Medicine

Tilyn N. DiGiacomo, PharmD, MSHIA, University of Alabama at Birmingham Hospital

Serena A. Dine, PharmD, FCCM, Indiana University Health

Sydney Dobson Kochheiser, PharmD, Indiana University Health

Cortney R. Dodson, PharmD, Prisma Health Richland

Kimberly L. Doerhoff, PharmD, MPA, SSM Health DePaul Hospital

Candiss Dominick, PharmD, University of Maryland Medical System

Logan J. Doriety, PharmD, McLeod Regional Medical Center

Sabrina Dunham, PharmD, University of Michigan Health

Kyle Dvoracek, PharmD, Forrest General Hospital

Emily Dye, PharmD, University of Alabama Birmingham

Megan E. Feeney, PharmD, Boston Medical Center

Christy C. Forehand, PharmD, FCCM, Piedmont Augusta; University of Georgia College of Pharmacy

Neal S. Fox, PharmD, Cedarville University School of Pharmacy

Amber D. Fraley, PharmD, Wellstar MCG Health/University of Georgia College of Pharmacy

Kayla Giang, PharmD, VA San Diego Healthcare System, UC San Diego Skaggs School of Pharmacy and Pharmaceutical Sciences, University of the Pacific Thomas J. Long School of Pharmacy

Caroline D. Gresham, PharmD, Piedmont Athens Regional

Kristin Griebe, PharmD, Henry Ford Hospital

Razelle Grimes, PharmD, HonorHealth Hospital and Medical Center

Ariel Haber, PharmD, Cleveland Clinic Florida

Rola A. Halabi, PharmD, MedStar Washington Hospital Center

Rachel A. Hall, PharmD, University of Maryland School of Pharmacy

Christian D. Hauser, PharmD, Indiana University Health Academic Health Center

Ashley Hawthorne, PharmD, Auburn University Harrison College of Pharmacy

Tanner L. Hedrick, PharmD, University of North Carolina at Chapel Hill - Eshelman School of Pharmacy

Brennan Herrmann, PharmD, University of Mississippi Medical Center

McKenzie J. Hodges, PharmD, Piedmont Columbus Regional Midtown

Alana K. Holliman, PharmD, University of Georgia College of Pharmacy

Caleb Hoover, PharmD, Premier Health Miami Valley Hospital

Erin Houry, PharmD, SSM Health Saint Louis University Hospital

Jessica Hu, PharmD, Ernest Mario School of Pharmacy at Rutgers, the State University of New Jersey

Wan-Ting Huang, PharmD, UC San Diego Health

Nicole E. Hume, PharmD, University of Kentucky

Kyle R. Humphreys, PharmD, UAB Medicine

Megan Ingebrigtson, PharmD, University of Michigan Health

Stephanie Janusz, PharmD, Miami Valley Hospital

Katelyn Jimison, PharmD, Atrium Health Wake Forest Baptist

Kayla E. John, PharmD, UNC Medical Center

Sara R. Jones, PharmD, University of Mississippi Medical Center

Nareg Kaltakdjian, PharmD, University of Georgia College of Pharmacy

Lauren C. Kennedy, PharmD, Barnes Jewish Hospital

Rebecca Kessinger, PharmD, Baylor St. Luke's Medical Center

Alley Killian, PharmD, Emory Healthcare

Natalie S. Kong, PharmD, Lankenau Medical Center

Christian L. Kressin, PharmD, University of Kentucky HealthCare

Christian Everett Kroll, PharmD, Mayo Clinic

Abigail M. Kurtz, PharmD, Sutter Memorial Medical Center

Megan C. Lail, PharmD, McLeod Regional Medical Center

Kaitlin M. Landolf, PharmD, University of Maryland School of Pharmacy

Ellen Lee, PharmD, Kadlec Regional Medical Center

Jennifer S. Lee, PharmD, Inova Fairfax Medical Campus

Kyla Leon, PharmD, University of Mississippi Medical Center

Matthew Li, PharmD, MHA, Westchester Medical Center

Mark J. Lin, PharmD, UCSF Benioff Children's Hospital Oakland

Nicole Lu, PharmD, Community Regional Medical Center

Whitney J. Ly, PharmD, University of North Carolina Medical Center

Katharine L. Madding, PharmD, Premier Health, Miami Valley Hospital

Olivia Marchionda, PharmD, Cleveland Clinic

Angelica Marques, PharmD, McLeod Regional Medical Center

Greg S. Martin, MD, MSc, Emory University School of Medicine

Whitney G. Mays, PharmD, University of Mississippi Medical Center

Bradford L. McDaniel, PharmD, MBA, Carilion Clinic

Allyson M. McIntire, PharmD, Franciscan Health Indianapolis

Brian P. McKinzie, PharmD, University of North Carolina

Mary McNeely, PharmD, Abbott Northwestern Hospital - Allina Health

Alyssa S. Meester, PharmD, The Ohio State University Wexner Medical Center

Arjay Mendoza, PharmD, VA San Diego Healthcare System

Kailey Meyer, PharmD, Avera Mckennan Hospital & University Health Center

Ashley E. Milkovits, PharmD, Carilion Clinic

James T. Miller, PharmD, University of Michigan Health

Christopher Miller, PharmD, MedStar Health

Keri L. Mills, PharmD, DCH Regional Medical Center

Corinne Murphy, PharmD, Piedmont Columbus Regional

David J. Murphy, MD, PhD, University of Colorado School of Medicine

Brian Murray, PharmD, University of Colorado Anschutz Medical Campus, Skaggs School of Pharmacy and Pharmaceutical Sciences

Neha D. Naik, PharmD, MBA, Emory Healthcare

Lama H. Nazer, PharmD, FCCM, King Hussein Cancer Center

Andrea M. Nei, PharmD, FCCM, Mayo Clinic

Jared Netley, PharmD, University of Wisconsin Hospitals and Clinics

Jennifer D. Nguyen, PharmD, UCSF Benioff Children's Hospital Oakland

Christian C. Nicolosi, PharmD, UMMC

Nicole M. Palm, PharmD, FCCM, Cleveland Clinic

Komal A. Pandya, PharmD, FCCM, University of Kentucky

Kristine A. Parbouni, PharmD, University of Maryland School of Pharmacy

Sara E. Parli, PharmD, UK HealthCare, University of Kentucky College of Pharmacy

Shyam Patel, PharmD, Boston Medical Center, Boston University School of Medicine

Akta S. Patel, PharmD, Mcleod Health

Kerilyn Petrucci, PharmD, UChicago Medicine

Brian Phan, PharmD, United Therapeutics Corporation

Stephanie T. Proctor, PharmD, Kadlec Regional Medical Center

Stephen H. Rappaport, PharmD, FCCM, University of Rochester Medical Center

Marianne Ray, PharmD, University of Mississippi College of Pharmacy

Paige Reese, PharmD, Wellstar Golisano’s Children’s Hospital of Georgia

Stephanee L. Rhoades, PharmD, ProMedica Toledo Hospital

Tim Robinson, PharmD, Wellstar MCG

Christine Rojas, BS, University of Maryland School of Pharmacy

Klayton M. Ryman, PharmD, UTSW

Alicia J. Sacco, PharmD, FCCM, Mayo Clinic

Mallorie N. Saling, PharmD, Jackson North Medical Center

Robert A. Sbertoli, PharmD, SSM Health Saint Louis University Hospital

Madeline A. Scarbrough, PharmD, UT Southwestern Medical Center

Addy E. Schoening, PharmD, Abbott Northwestern Hospital - part of Allina Health

Janet Shin, PharmD, UCSF Benioff Children's Hospital Oakland

Zachary R. Smith, PharmD, FCCP, FCCM, Henry Ford Hospital

Brooke A. Smith, PharmD, Wellstar MCG

Brandon Smith, PharmD, Prisma Health Richland Hospital

Maya Smith, PharmD, Luminis Health Anne Arundel Medical Center

Alyssa Sonchaiwanich, PharmD, Mayo Clinic Rochester

Katherine Spezzano, PharmD, MBA, University of Kentucky HealthCare

Gillian Steiger, PharmD, Massachusetts General Hospital

Donna A. Steinbacher, PharmD, UNC Health

Joseph A. Stoldt, PharmD, IU Health

Margaret A. Street, PharmD, Baylor St. Luke's Medical Center, Houston, TX

Maria K. Stubbs, BS Pharm, VA San Diego Healthcare System; UC San Diego Skaggs School of Pharmacy and Pharmaceutical Sciences; University of the Pacific Thomas J. Long School of Pharmacy

Jennifer Tawwater, PharmD, UT Southwestern Medical Center, Dallas, TX

Ashley N. Taylor, PharmD, Wellstar MCG Health Medical Center

Dakota Taylor, PharmD, University of Mississippi Medical Center

Tori Thompson, PharmD, MercyOne Des Moines Medical Center

Kailee A. Toews, PharmD, Community Regional Medical Center

Tu-Trinh T. Tran, PharmD

Abby Tyson, PharmD, OhioHealth Riverside Methodist Hospital

Sandra A. Valencia, PharmD, Providence Sacred Heart Medical Center

Meghna Vallabh, PharmD, Baylor St. Luke's Medical Center

Storm A. Van Wey, PharmD, Indiana University School of Medicine, Butler University College of Pharmacy & Health Sciences, Purdue University College of Pharmacy

Beth E. Varnes, PharmD, UAB Hospital

Vanessa C. Velazco, PharmD, Cleveland Clinic Florida

Arianna J. Vidger, PharmD, Indiana University Health

Dalena Vo, PharmD, Indiana University Health, Academic Health Center

Ann Vu, PharmD, Community Regional Medical Center

Stephanie C. Waldrep, PharmD, University of Alabama at Birmingham

Jessica A. Ward, PharmD, Cleveland Clinic

Nathaniel B. Wayne, PharmD, Wellstar MCG Health, University of Georgia College of Pharmacy

Lori Wetmore, PharmD, Shenandoah University

Jessica A. Whitten, PharmD, Eskenazi Health

Alexandra M. Wiegand, PharmD, UK HealthCare

Sarah K. Williford, PharmD, University of North Carolina Health

Sharon Wilson, PharmD, University of Maryland Medical Center

Kevin M. Wohlfarth, PharmD, ProMedica Toledo Hospital

Douglas R. Wylie, PharmD, University of Alabama at Birmingham

Siu Yan A. Yeung, PharmD, University of Maryland Medical Center

Jae H. Yook, PharmD, UGA College of Pharmacy

Connie H. Yoon, PharmD, OhioHealth Riverside Methodist Hospital

### **List of Acknowledgements**

REDCap Consortium at Vanderbilt

Pharmaceutical Research Computing (PRC) center at University of Maryland School of Pharmacy

Society of Critical Care Medicine (SCCM)

American College of Clinical Pharmacy (ACCP)

Diana Aguilar

Ghadah Alajmi

Julia Alexander

Emily Austin

Cesar Bejarano-Garcia

Andrew T. Bennett

Jessica M. Biggs

Mary Blair

Kaitlin M. Blotske

James Braun

Nicholas Bravo

Garrett Brown

Jennifer Bui

Joshua Campbell

Carlette Cavenaugh

Michael E. Chao

Sarah Chiu

Patrick Costello

Samantha Delibert

Sam Dewitt

Megan Dorsey

Courtney Feagin

Shelby Fideler

Samantha L. Gauthier

Emily George

Kristen Giles

Alexis Glenn

Renae Gozelski

Aileen Gregorio-Corallo

Liana Ha

Andrea Hankins

Gresham Hindman

Elizabeth Hodges

Cory Johnson

Kevin Josey

Lama Kanawati

Nadine Kanyana

Jana L. Kelly

Tara Kennell

Alexa Luboff

Isabel Mangaoang

Carolyn Martz

Taylor McCart

Jamie McCarthy

Ana McLean

Stephanie Millan

Emily Miller

Makenna Moll

Peter Moran

Rebecca Morgan

Amoreena Most

Zach Muller

Nicole Newton

Kelly T. Nguyen

Justin Petrovic

Inna Perinskaya

Nicole Pfeffer

Kara E. Phillips

Leslie Phillips

Lisa Pickmans

Laura Provost

Caitlin J. Quibeuf

Marinna Raqueno

Jenna Schwartz

Shaleen Singh

Jillian K. Songstad

Kelcy Sorsensen

Inderpal Srai

Melissa Sterling

Samori Swygert

Kari Taggart

Farrah Tavakoli

Melissa Thompson Bastin

Dalena Vo

Noelle Vo

Todd Walroth

Hailey Wang

Brian Watson

Ashley Wischmeyer

Laura Witt

Shelby Young

Denisse Garcia Zavala

Kara Zacholski

Qingrong Laura Zhang

### **List of Disclosures**

Authors with conflicts of interest are listed below. If an author is not listed, they reported no conflicts of interest.

Marisha Burden- Dr. Burden reports funding from the Agency for Healthcare Research and Quality, the National Institute for Occupational Health and Safety, University of Colorado Innovations digiSPARK award, and the American Medical Association not related to this work. Dr. Burden contributed to the development of GrittyWork, a digital workforce application, and a registered trademark of the University of Colorado, not related to this work. Dr. Burden reports honorarium from Med-IQ not related to this work.

Ashley Hawthorne- Speaker’s Bureau for Vericel Corporation

Christy Forehand- sits on the Board of Pharmacy Specialties Critical Care Specialty Council (volunteer, unpaid position)

Susan Smith- sits on the Advisory Board for Amneal Pharmaceuticals, Inc.

Douglas Wylie- employed by Chiesi USA

### **Reporting of Observational Studies in Epidemiology (STROBE) Checklist**

|  | **Item No** | **Recommendation** | **Page No** |
| --- | --- | --- | --- |
| **Title and abstract** | 1 | (*a*) Indicate the study’s design with a commonly used term in the title or the abstract | 1 |
|  |  | (*b*) Provide in the abstract an informative and balanced summary of what was done and what was found | 2 |
| **Introduction** | | | |
| Background/rationale | 2 | Explain the scientific background and rationale for the investigation being reported | 3 |
| Objectives | 3 | State specific objectives, including any prespecified hypotheses | 3 |
| **Methods** | | | |
| Study design | 4 | Present key elements of study design early in the paper | 4 |
| Setting | 5 | Describe the setting, locations, and relevant dates, including periods of recruitment, exposure, follow-up, and data collection | 4 |
| Participants | 6 | (*a*) Give the eligibility criteria, and the sources and methods of selection of participants. Describe methods of follow-up | 4 |
|  |  | (*b*) For matched studies, give matching criteria and number of exposed and unexposed |  |
| Variables | 7 | Clearly define all outcomes, exposures, predictors, potential confounders, and effect modifiers. Give diagnostic criteria, if applicable | 4 |
| Data sources/ measurement | 8* | For each variable of interest, give sources of data and details of methods of assessment (measurement). Describe comparability of assessment methods if there is more than one group | 4 |
| Bias | 9 | Describe any efforts to address potential sources of bias | 4 |
| Study size | 10 | Explain how the study size was arrived at | 4 |
| Quantitative variables | 11 | Explain how quantitative variables were handled in the analyses. If applicable, describe which groupings were chosen and why | 4 |
| Statistical methods | 12 | (*a*) Describe all statistical methods, including those used to control for confounding | 4 |
|  |  | (*b*) Describe any methods used to examine subgroups and interactions |  |
|  |  | (*c*) Explain how missing data were addressed | 4 |
|  |  | (*d*) If applicable, explain how loss to follow-up was addressed |  |
|  |  | (*e*) Describe any sensitivity analyses |  |
| **Results** | | |  |
| Participants | 13* | (a) Report numbers of individuals at each stage of study—eg numbers potentially eligible, examined for eligibility, confirmed eligible, included in the study, completing follow-up, and analysed | 6 |
|  |  | (b) Give reasons for non-participation at each stage | 11 |
|  |  | (c) Consider use of a flow diagram | 11 |
| Descriptive data | 14* | (a) Give characteristics of study participants (eg demographic, clinical, social) and information on exposures and potential confounders | 6 |
|  |  | (b) Indicate number of participants with missing data for each variable of interest | 11 |
|  |  | (c) Summarise follow-up time (eg, average and total amount) |  |
| Outcome data | 15* | Report numbers of outcome events or summary measures over time | 6 |
| Main results | 16 | (*a*) Give unadjusted estimates and, if applicable, confounder-adjusted estimates and their precision (eg, 95% confidence interval). Make clear which confounders were adjusted for and why they were included | 6 |
|  |  | (*b*) Report category boundaries when continuous variables were categorized |  |
|  |  | (*c*) If relevant, consider translating estimates of relative risk into absolute risk for a meaningful time period |  |
| Other analyses | 17 | Report other analyses done—eg analyses of subgroups and interactions, and sensitivity analyses |  |
| **Discussion** | | | |
| Key results | 18 | Summarise key results with reference to study objectives | 8 |
| Limitations | 19 | Discuss limitations of the study, taking into account sources of potential bias or imprecision. Discuss both direction and magnitude of any potential bias | 9 |
| Interpretation | 20 | Give a cautious overall interpretation of results considering objectives, limitations, multiplicity of analyses, results from similar studies, and other relevant evidence | 8 |
| Generalisability | 21 | Discuss the generalisability (external validity) of the study results | 9 |
| **Other information** | | | |
| Funding | 22 | Give the source of funding and the role of the funders for the present study and, if applicable, for the original study on which the present article is based | ii |

*Give information separately for exposed and unexposed groups.

**Note:** An Explanation and Elaboration article discusses each checklist item and gives methodological background and published examples of transparent reporting. The STROBE checklist is best used in conjunction with this article (freely available on the Web sites of PLoS Medicine at http://www.plosmedicine.org/, Annals of Internal Medicine at http://www.annals.org/, and Epidemiology at http://www.epidem.com/). Information on the STROBE Initiative is available at http://www.strobe-statement.org.

### **Supplemental Figure 1. Correlation Matrix of Predictors Included in the Mortality Model**

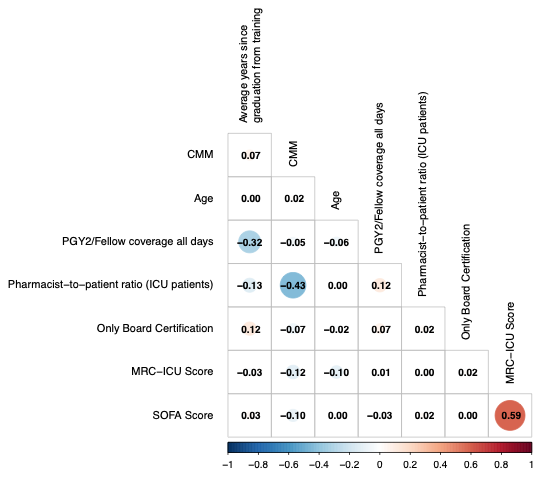

CMM: comprehensive medication management; ICU: intensive care unit; MRC-ICU: Medication regimen complexity-Intensive care unit; PGY: Post-graduate year; SOFA: Sequential Organ Failure Assessment

### **Supplemental Table 1. Variance Inflation Factors (VIF) for Predictors Included in the Multivariable Model**

| **Variable** | **VIF** |
| --- | --- |
| MRC-ICU Score | 1.569354 |
| SOFA Score | 1.541952 |
| CMM delivered on interprofessional rounds | 1.257878 |
| Pharmacist-to-ICU patient ratio | 1.256689 |
| Average years since graduation from training | 1.153549 |
| Coverage by a pharmacist who completed a PGY2 or fellowship every day of ICU admission | 1.136806 |
| BCP | 1.031749 |
| Age | 1.021898 |

BCP: Board-certified pharmacist; CMM: comprehensive medication management; ICU: intensive care unit; MRC-ICU: Medication regimen complexity-Intensive care unit; PGY: Post-graduate year; SOFA: Sequential Organ Failure Assessment

### **Supplemental Table 2. Missing Data**

| **Variable** | **All Patients (N=20,537)** | **Non-BCP (N=2,097)** | **BCP**  **(N=18,440)** |
| --- | --- | --- | --- |
| **CCP-to-ICU-patient ratio** | 0 (0) | 0 (0) | 0 (0) |
| **Age** | 0 (0) | 0 (0) | 0 (0) |
| **Sex** | 0 (0) | 0 (0) | 0 (0) |
| **SOFA Score*** | 0 (0) | 0 (0) | 0 (0) |
| **MRC-ICU Score*** | 0 (0) | 0 (0) | 0 (0) |
| **ICU Admission Day of the Week** | 0 (0) | 0 (0) | 0 (0) |
| **ICU Type** | 0 (0) | 0 (0) | 0 (0) |
| **Institution Type** | 0 (0) | 0 (0) | 0 (0) |
| **RN-to-ICU-patient ratio** | 0 (0) | 0 (0) | 0 (0) |
| **Coverage by a pharmacist who completed a PGY2 or fellowship every day of ICU admission** | 0 (0) | 0 (0) | 0 (0) |
| **CMM delivered on interprofessional rounds every day of ICU** | 0 (0) | 0 (0) | 0 (0) |
| **CCP coverage 1st 24h of ICU stay** | 6 (<0.1) | 1 (<0.1) | 5 (<0.1) |
| **Average years since graduation from training** | 0 (0) | 0 (0) | 0 (0) |
| **Mechanical ventilation** | 0 (0) | 0 (0) | 0 (0) |
| **Mechanical circulatory support** | 1 (<0.1) | 0 (0) | 1 (<0.1) |
| **Dialysis** | 1 (<0.1) | 0 (0) | 1 (<0.1) |
| **Mortality** | 0 (0) | 0 (0) | 0 (0) |
| **Hospital Length of Stay** | 1(<0.1) | 1(<0.1) | 0 |
| **ICU Length of Stay** | 0 (0) | 0 (0) | 0 (0) |

All data presented as count (percentage)

BCP: Board-certified pharmacist; SOFA: Sequential Organ Failure Assessment; MRC-ICU: Medication regimen complexity-Intensive care unit; ICU: Intensive care unit; RN: Registered Nurse; CMM: Comprehensive Medication Management; PGY: Post-graduate year

*worst score during the first 24 hours of ICU stay
